# Estimating the Growth Rate and Doubling Time for Short-Term Prediction and Monitoring Trend During the COVID-19 Pandemic with a SAS Macro

**DOI:** 10.1101/2020.04.08.20057943

**Authors:** Stanley Xu, Christina Clarke, Susan Shetterly, Komal Narwaney

## Abstract

Coronavirus disease (COVID-19) has spread around the world causing tremendous stress to the US health care system. Knowing the trend of the COVID-19 pandemic is critical for the federal and local governments and health care system to prepare plans. Our aim was to develop an approach and create a SAS macro to estimate the growth rate and doubling time in days if growth rate is positive or half time in days if growth rate is negative. We fit a series of growth curves using a rolling approach. This approach was applied to the hospitalization data of Colorado State during March 13^th^ and April 13^th^. The growth rate was 0.18 (95% CI=(0.11, 0.24)) and the doubling time was 5 days (95% CI= (4, 7)) for the period of March 13^th^-March 19^th^; the growth rate reached to the minimum −0.19 (95% CI= (−0.29, −0.10)) and the half time was 4 days (95% CI= (2, 6)) for the period of April 2^nd^ – April 8^th^. This approach can be used for regional short-term prediction and monitoring the regional trend of the COVID-19 pandemic.

## 1. BACKGROUND

In December 2019, an outbreak of coronavirus disease (COVID-19) caused by the novel coronavirus (SARS-CoV-2) began in Wuhan, China and has now spread across the world [1,2]. In the United States, the cumulative number of identified COVID-19 cases was 186,101 as of March 31st, 2020; among the identified cases, 3603 died [3]. To slow the spread of COVID-19, federal and local governments have issued mitigation measures such as case isolation, quarantine, school closures and closing non-essential businesses. The COVID-19 pandemic imposes tremendous challenges to the US health care system, particularly given concerns that the need for hospital beds and ICU beds could exceed capacity [4-6]. Predicting the future numbers of COVID-19 cases and healthcare utilization is critical for governments and health care systems preparation plans [4,6,7]. Two useful and critical quantities for prediction are the growth rate [8] and the doubling time of number of events [9]. The growth rate is the percent change of daily events (e.g, COVID-19 cases, number of patients hospitalized or number of deaths). The doubling time is the length of time required to double the number of daily events.

Our goal was to develop an approach and create a SAS macro using observed data to estimate the growth rate and doubling time in days for short-term prediction.

## 2. METHODS

### 2.1 A rolling growth curve approach (RGCA)

In the United States, there are several barriers for testing people for COVID-19 such as shortages of swabs and testing kits and restrictions on who should get tested.

Therefore, the number of COVID-19 cases is often under-identified and under-reported. However, the number of hospitalized COVID-19 patients (hospitalizations) and number of deaths due to COVID-19 are more reliable than the reported number of COVID-19 cases [10]. In this paper, we used the number of daily hospitalized COVID-19 patients to calculate the growth rate and doubling time in days.

We assumed a growth curve of daily hospitalizations over a period of n days from day t (start day) to day (*t* + *n* − 1). Let *y*_(*t*+*J* − 1)_ denote the daily hospitalizations at day (*t* + − 1), 1 ≤*j* ≤ n. Based on the growth model, we have

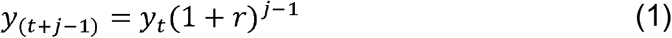

where y_t_ is the number of hospitalizations at the start day *t*; *r* is the growth rate. When the growth rate *r* > 0, the number of daily hospitalizations increases. For example, if *r* =0.4, the growth rate of hospitalizations is 40% more for each day. When growth rate *r* = 0, the number of daily hospitalizations has no change. When growth rate *r* < 0, the number of daily hospitalizations declines. When the number of hospitalizations doubles at *j* = *D*, that is *y*_*(t+D − 1)*_ = 2*y*_*t*_, we have,

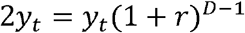

Further, it can be shown that

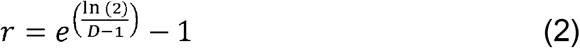

We fit two models: a) using equation (1) which estimates the growth rate r; b) using equation (1) with *r* substituted with e 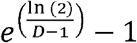 from equation (2). The second model estimates the doubling time in days a, meaning that it takes D days from the start day *t* for the number of daily hospitalizations to double. We used SAS PROC NLIN [11] to fit these two nonlinear models. Note that equation (2) is valid for *r* > 0. When *r* < 0, one can use 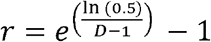 the estimated a represents the days required to reduce the number of hospitalizations by half (half time).

Because the growth rate and doubling time may change over time, we used a rolling growth curve approach (RGCA). For example, we set the length of the period to be 7 days (n = 7 days). We estimated the growth rate and the doubling time in days for the following periods for hospitalization data from Colorado State from March 13th – April 13th [12]: March 13^th^-19^th^,14^th^-20^th^, 15^th^-21^st^,…, April 7^th^-April 13^th^

### 2.2 Short-term prediction

The estimated growth rate from the last period of the RGCA approach (e.g., April 7^th^-April 13^th^) can be used for future short-term prediction of hospitalizations. Let *k* denote the last day of the last period, *y*_*k*_ is the number of hospitalizations on this day. For the Colorado hospitalization data in this analysis, *k* is April 13^th^, *y*_k_ = 36. Let *m* denote the date after date *k*, then the predicted *y*_*m*_is

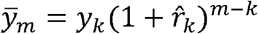

Where 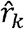 is the estimated growth rate from the last period. As the growth rate changes over time, the prediction is only appropriate for short-term prediction (e.g., within 7 days) and updated growth rates should be used.

## 3. RESULTS

We estimated a series of growth rates using RGCA with a length of 7 days. The estimated growth rates and 95% CIs were plotted over time using the mid-day of a 7 day period (Figure 1). The growth rate peaked with a value of 56.2% at the mid-day of March 18^th^ for the period March 15^th^ and March 21^st^. Between March 18^th^ and April 1^st^, although the growth rate continuously decreased, the daily number of hospitalizations increased because of positive growth rates. We started to observe negative growth rates after April 1^st^, except for a positive growth rate on April 9^th^. The growth rate reached its minimum at the mid-day of April 5^th^ (period April 2^nd^-April 8^th^) with a value of −19.2%. The growth rate then increased after April 5^th^. Note that a negative growth rate represents a reduction in number of hospitalizations.

**Figure 1.**
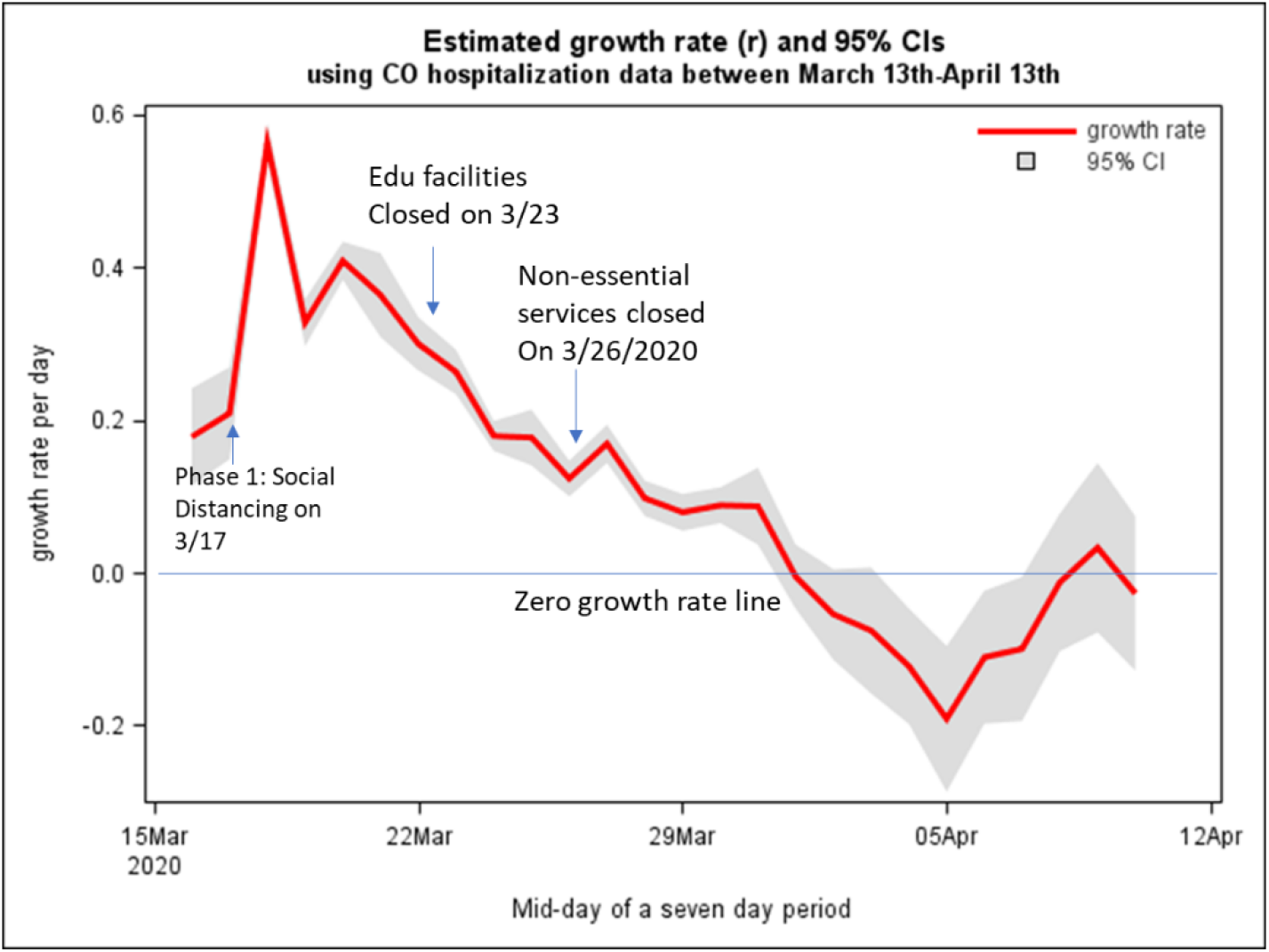
Estimated growth rate with 95% CIs over time using hospitalization data from Colorado State.

The doubling time (growth rate>0) and half time (growth rate<0) in days over time are displayed in Figure 2. Before April 1^st^, the y-axis represents the doubling time in days because of positive growth rates. After April 1^st^, except for a positive growth rate on April 9^th^, the y-axis represents a half-time because of negative growth rates. On April 1^st^, the reduction rate was very small (0.5%) which resulted in a high half-time, 128 days with very wide 95% CIs (not shown in the figure). On April 8^th^, there was a small reduction rate (1.2%) resulting in 55 days of half time.

**Figure 2.**
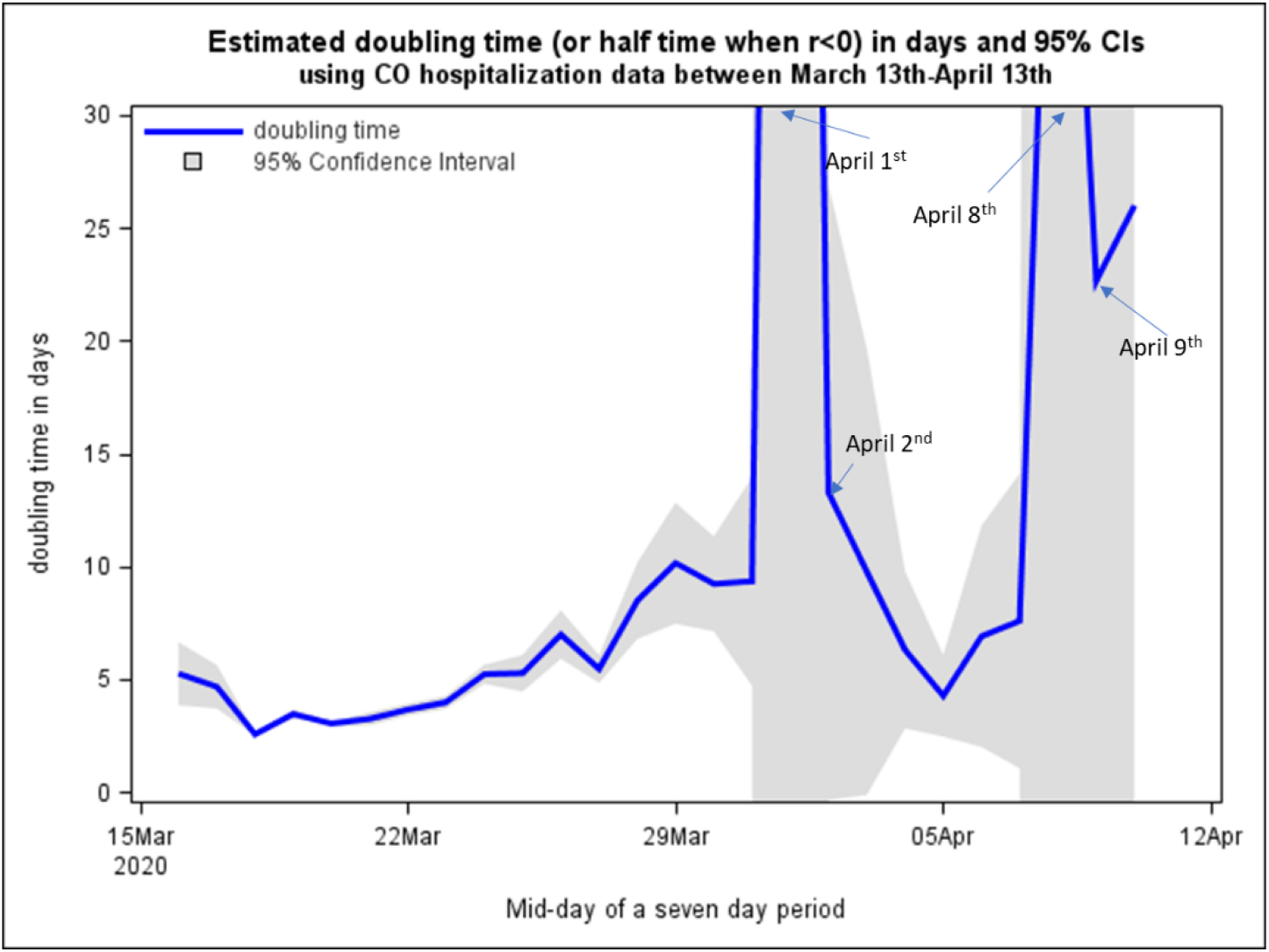
Estimated doubling (or half) time in days with 95% CIs over time using hospitalization data from Colorado State.

Using the estimated growth rate from the last period April 7^th^-April 13^th^, rk = 0.027, the predicted numbers of daily hospitalizations for April 14^th^ and 15^th^ were 35 and 34, respectively. SAS programs are available for conducting these analyses (Appendix A and Appendix B).

## 4. DISCUSSION

These models can be similarly applied to death data if they are available and not sparse. When COVID-19 testing is widely available to the public and the number of COVID-19 testing is less selective, these models can also be used to directly estimate the growth rate and the doubling time for COVID-19 cases. Due to a lag in reporting hospitalization, it is recommended to exclude the recent 1-2 days’ hospitalization data in fitting the growth curves. This paper illustrates that hospitalization data can be used to estimate the growth rate and doubling (or half) time to aid predicting future hospitalizations, deaths and COVID-19 cases. Because a series of growth curves were fit, the RGCA approach can also be used for real-time monitoring of the epidemic trend as shown in Figure 1.

Colorado state issued three social distancing guidelines: a voluntary social distancing on March 17^th^, closing educational facilities on March 23^rd^, and closing non-essential services on March 26^th^ (Figure 1). It takes some time (e.g., 2 weeks) for these mitigation measures to have impact. Although the effectiveness of these mitigation measures has not been investigated formally, it is believed that they helped to slow the spread of COVID-19 and reduced number of hospitalizations and death in Colorado and across the United States.

## Data Availability

all data available the provided website

https://coronavirus.1point3acres.com/en

## Acknowledgements

This research was supported by the Institute for Health Research, Kaiser Permanente Colorado. Xu was also supported by NIH/NCRR Colorado CTSI Grant Number UL1 RR025780.

## APPENDIX A

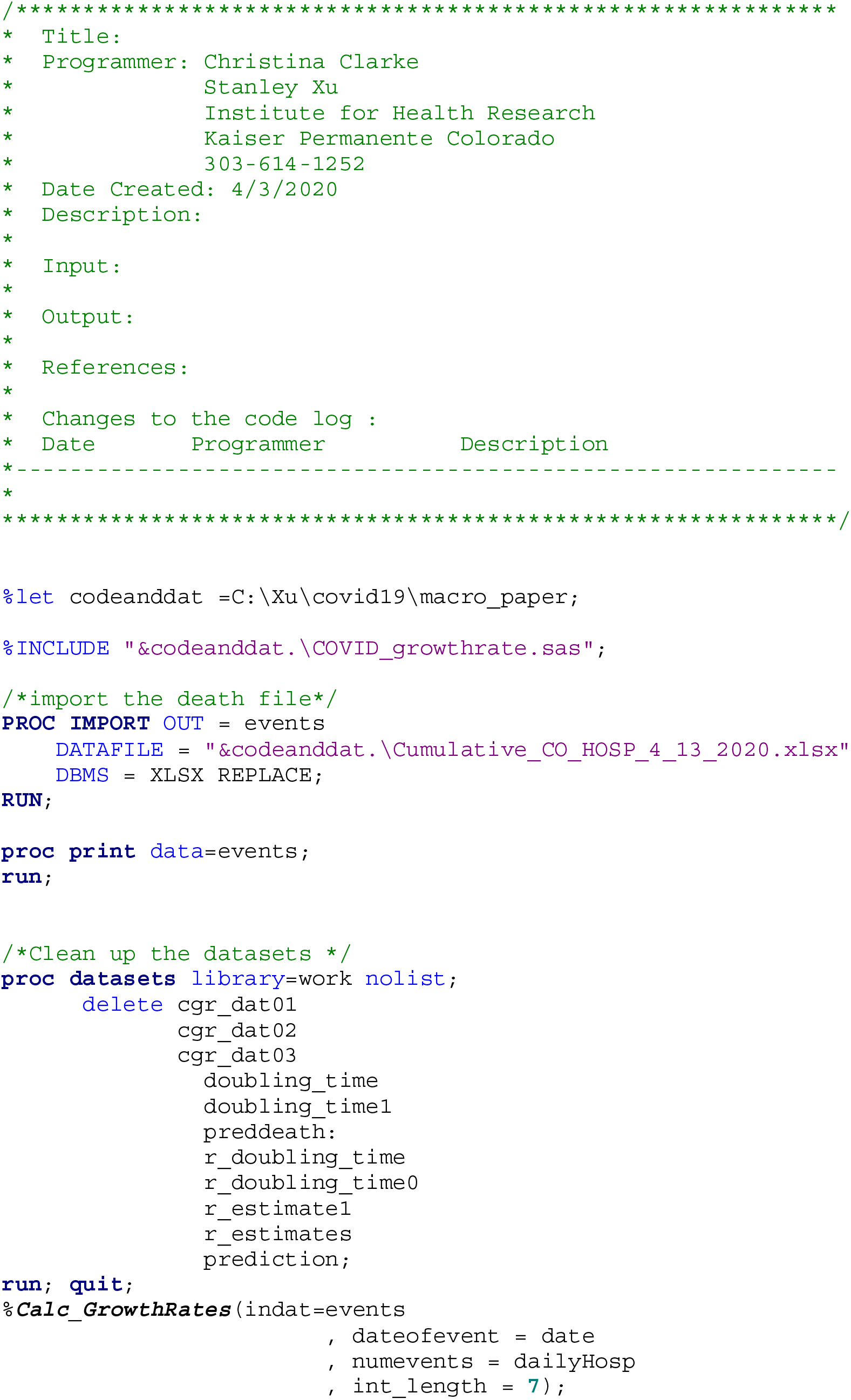
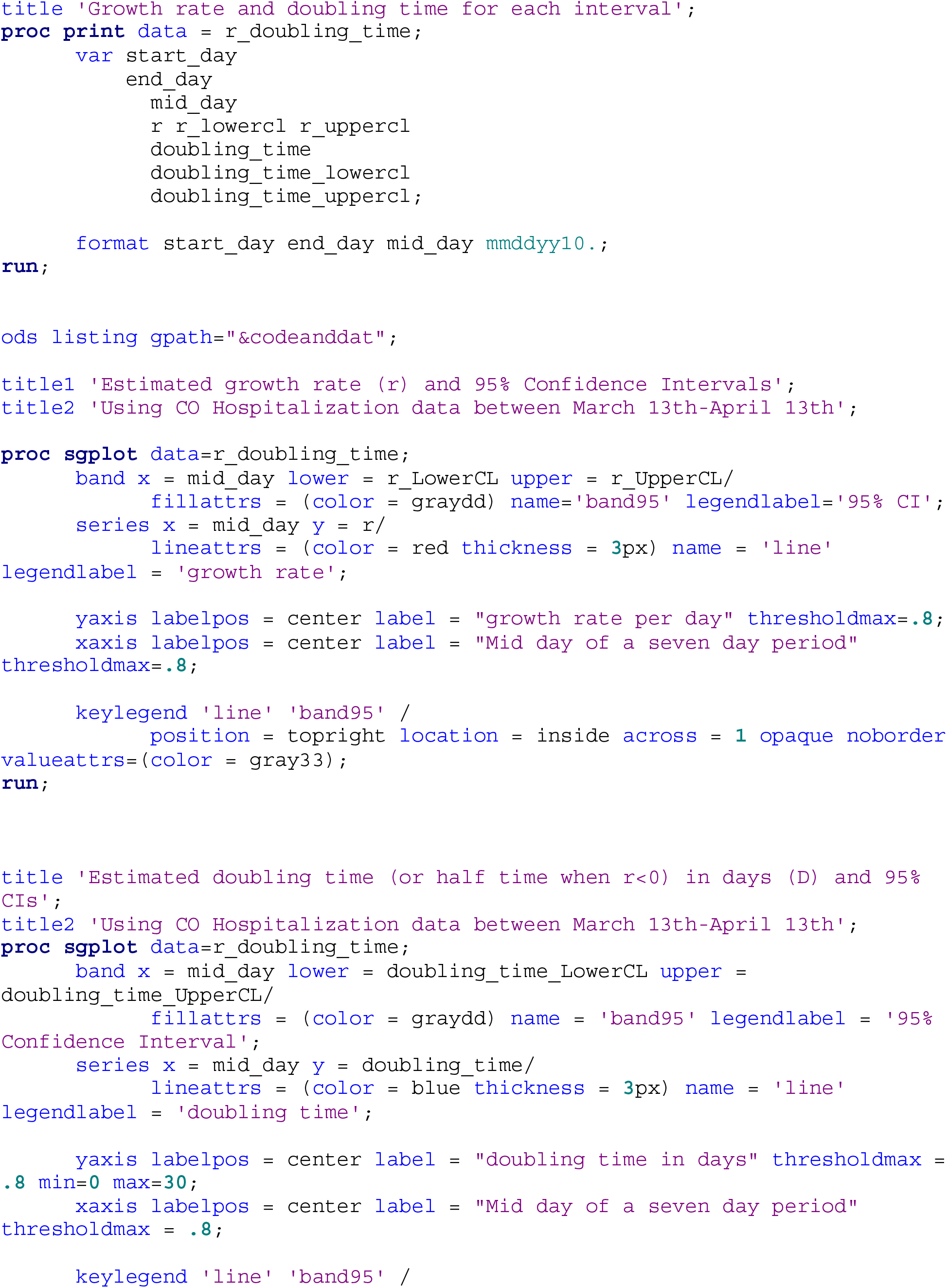
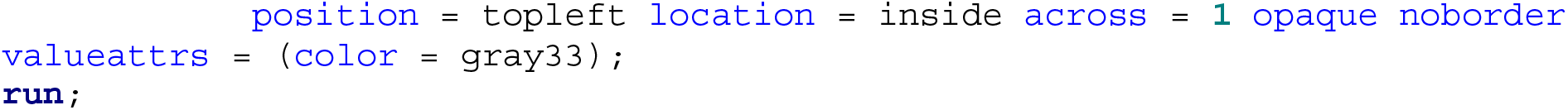

## APPENDIX B

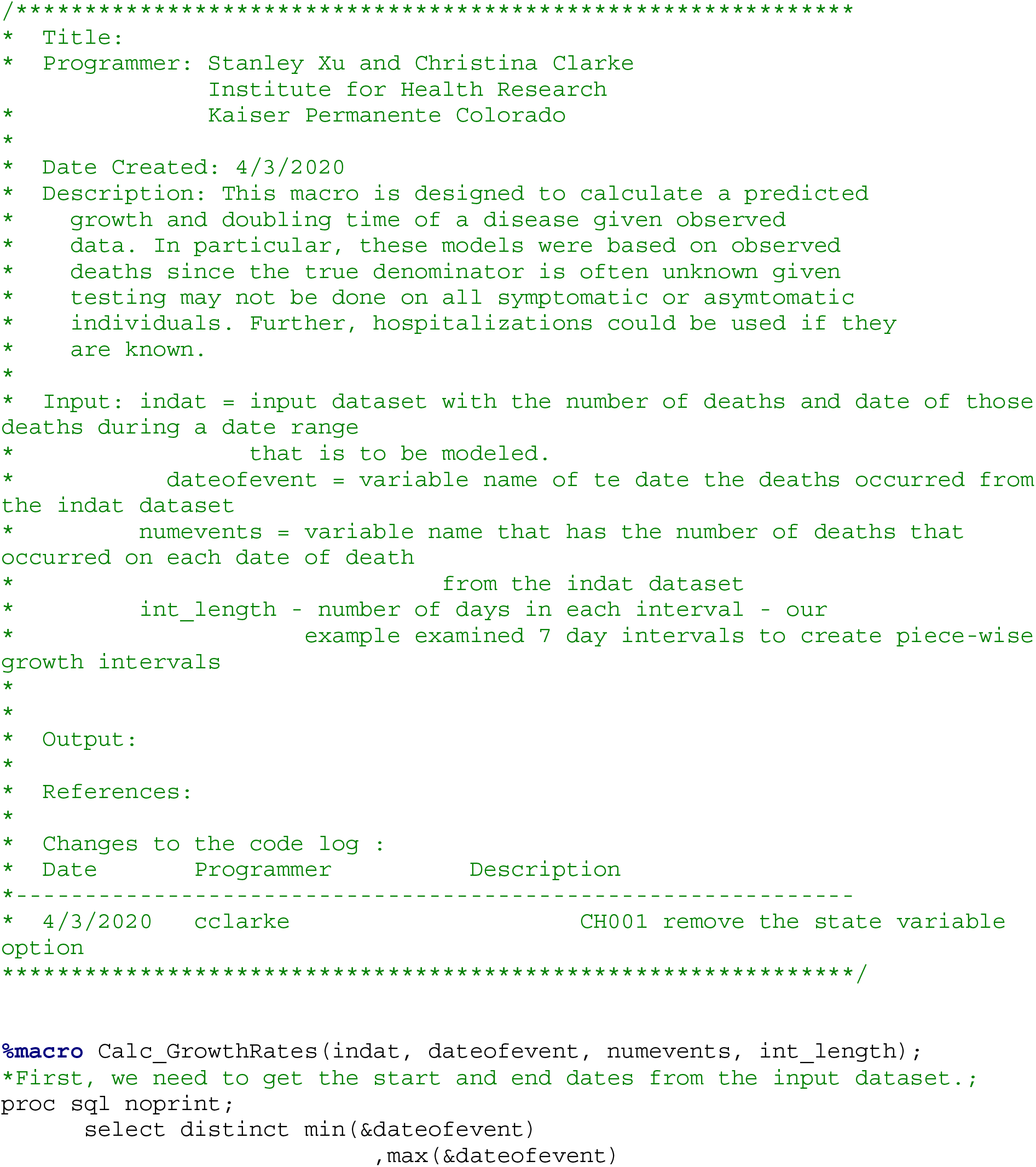
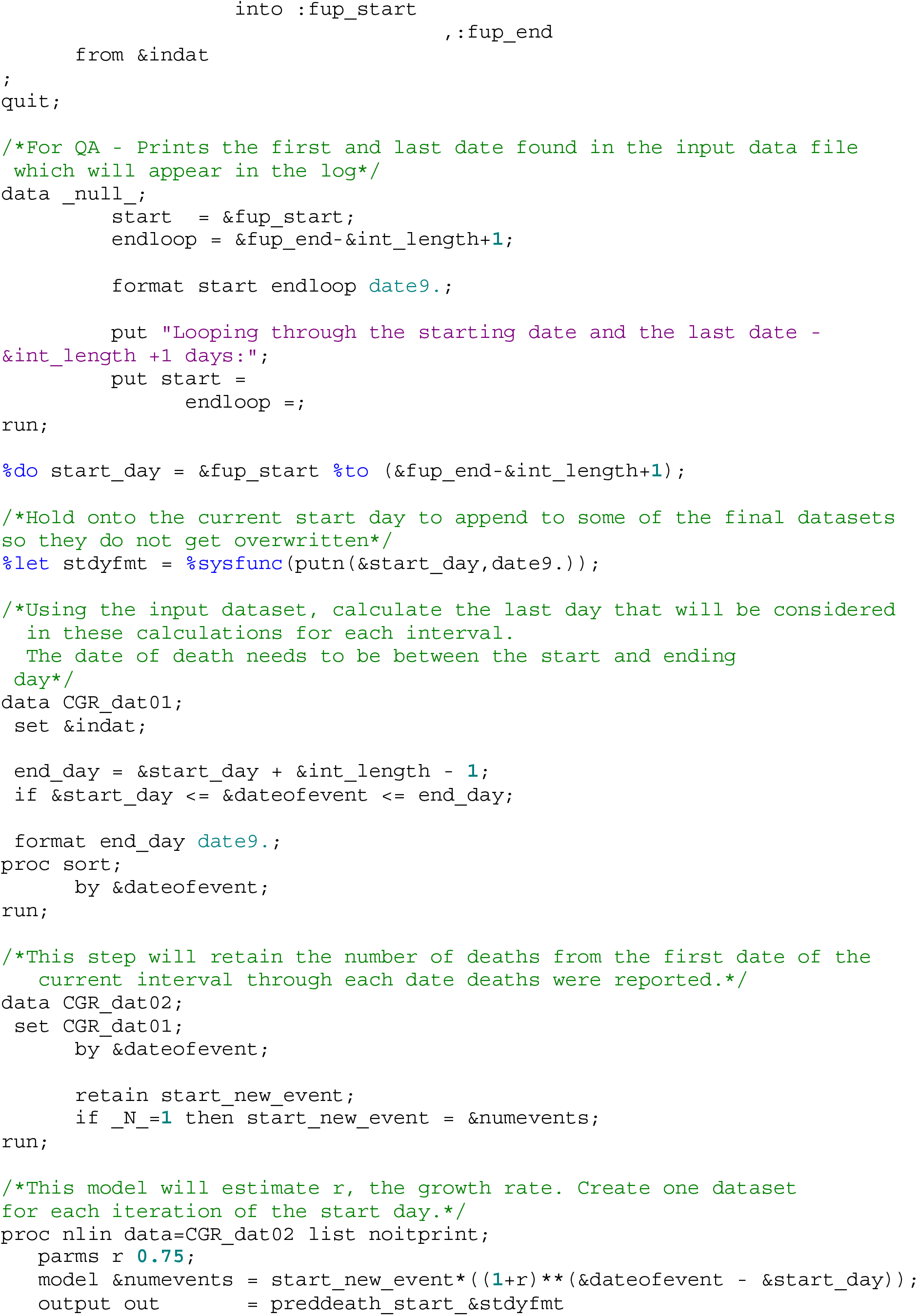
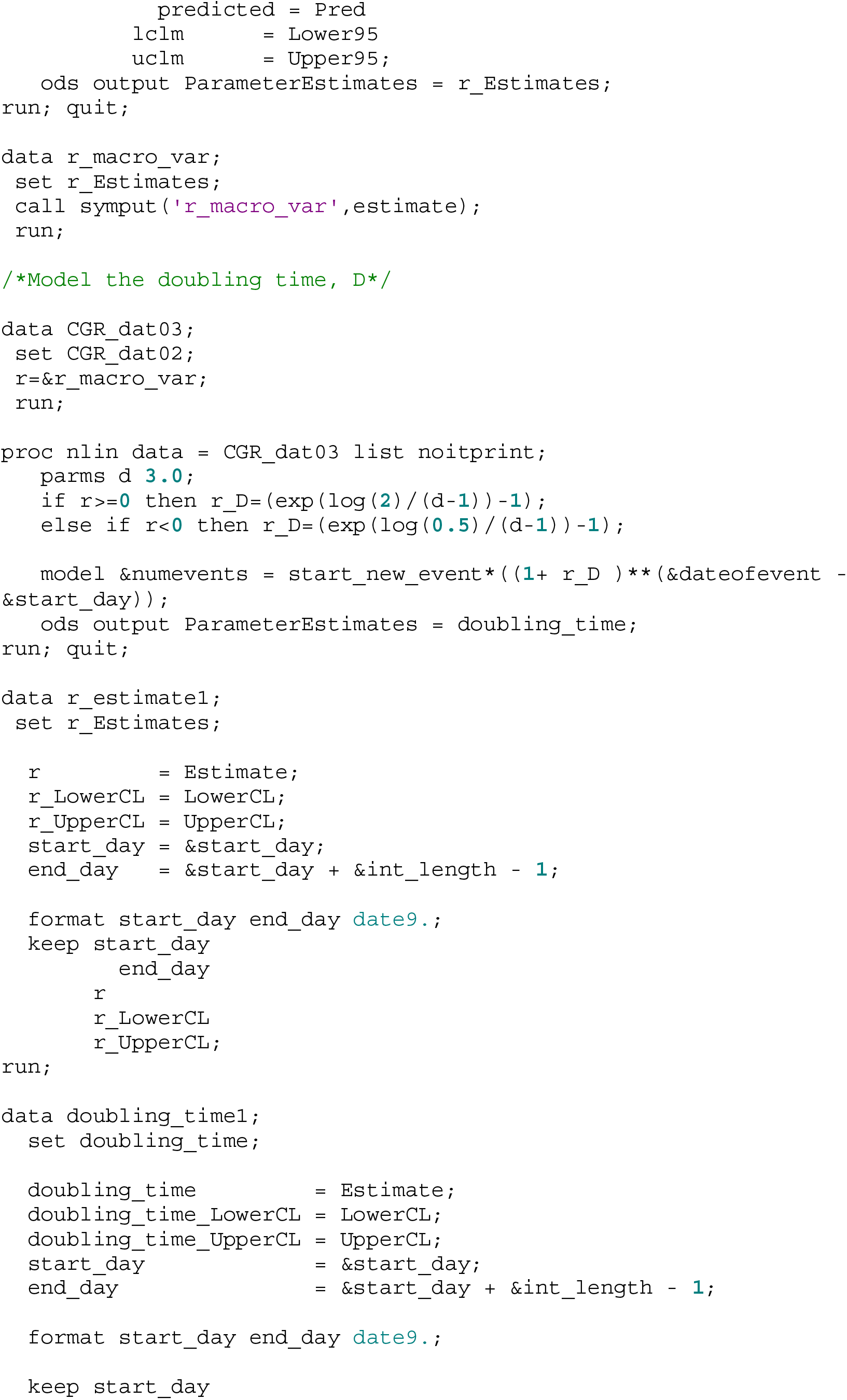
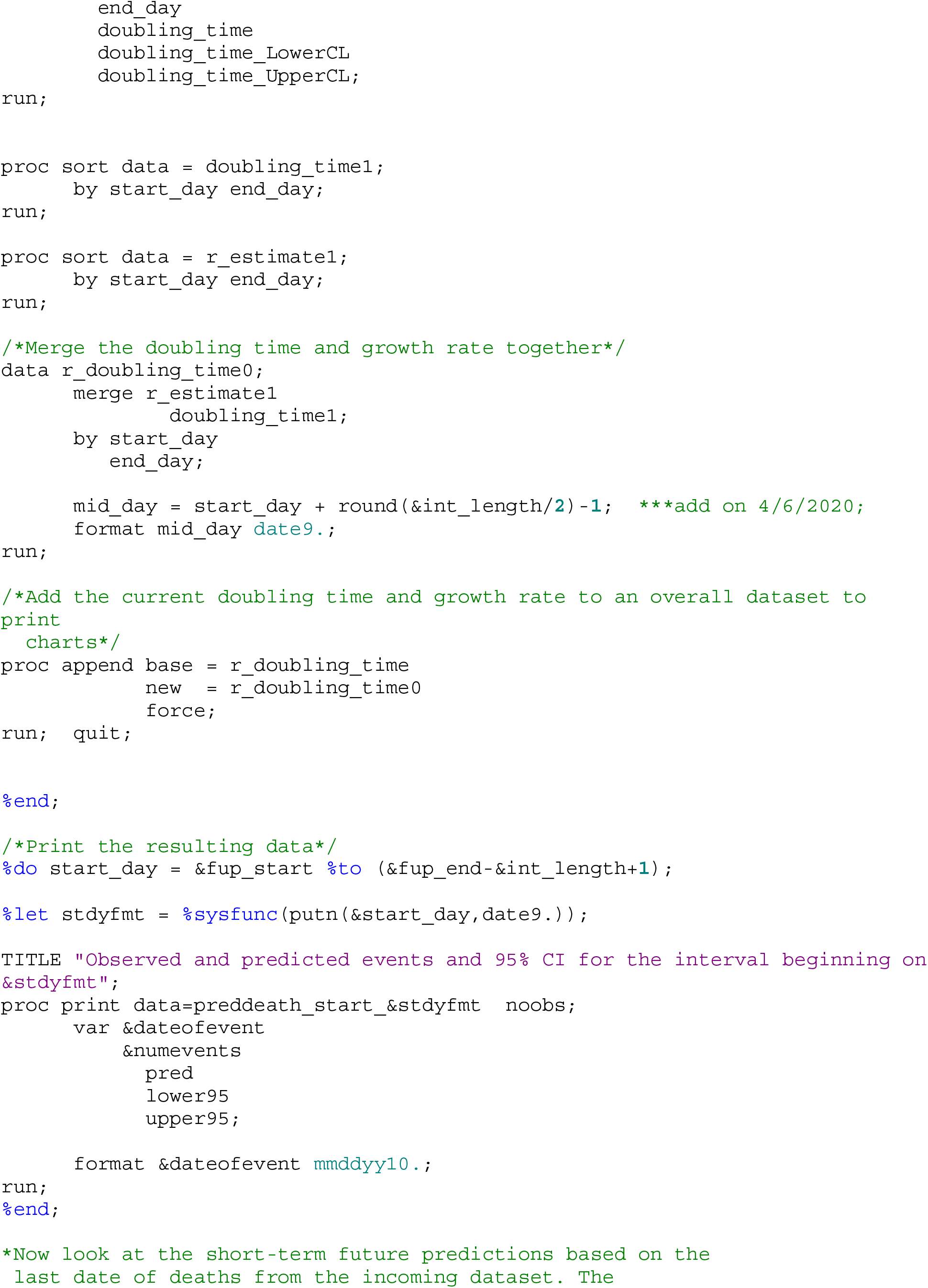
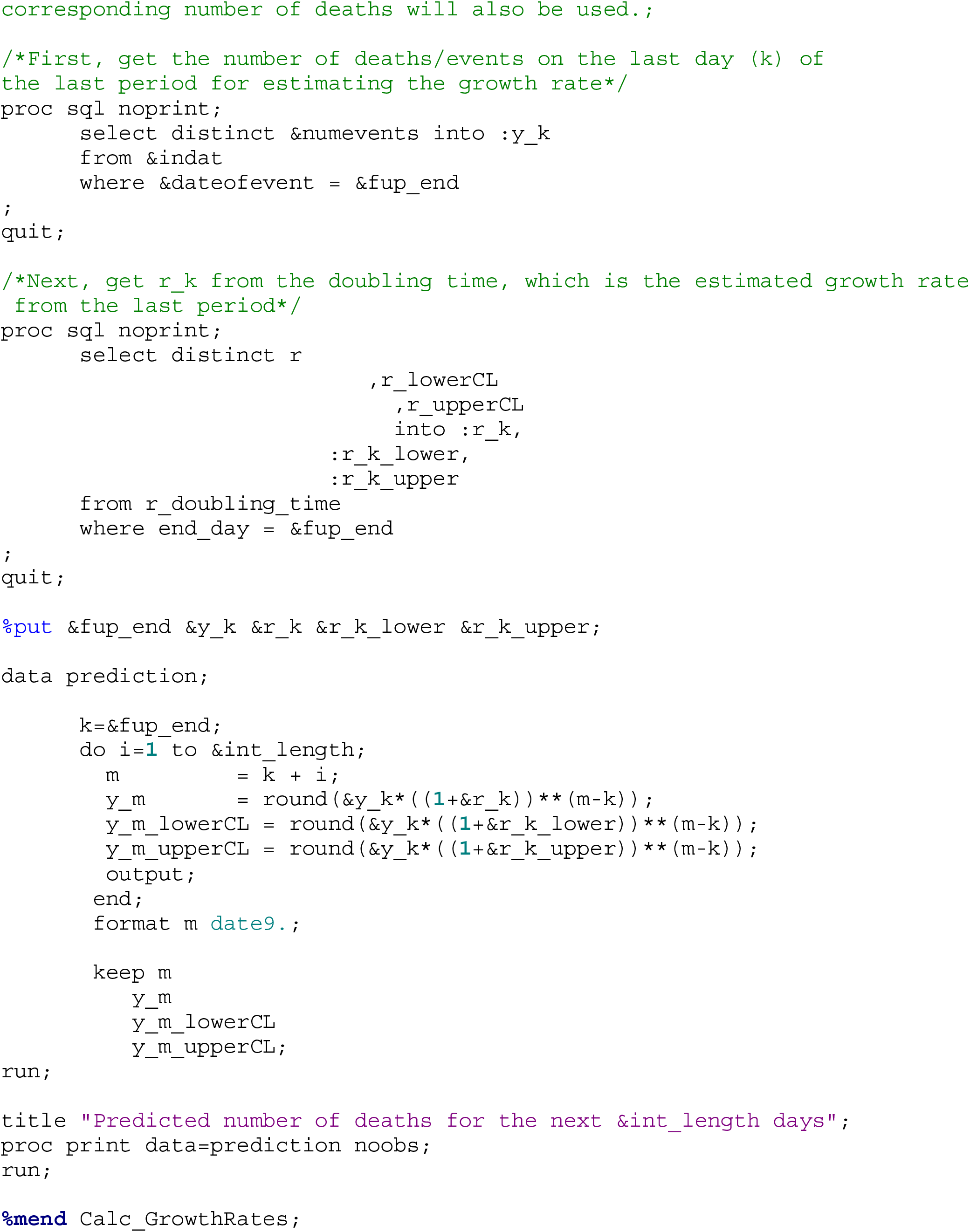

## REFERENCES

1. CDC. 2019 Novel Coronavirus, Wuhan, China. Available at https://www.cdc.gov/coronavirus/2019-ncov/

2. WHO. WHO Director-General’s opening remarks at the media briefing on COVID-19 – 11 March 2020

3. CDC. https://www.cdc.gov/coronavirus/2019-ncov/cases-updates/cases-in-us.html

4. IHME COVID-19 health service utilization forecasting team. Forecasting COVID-19 impact on hospital bed-days, ICU-days, ventilator days and deaths by US state in the next 4 months. MedRxiv. 26 March 2020. doi:10.1101/2020.03.27.20043752.

5. Ferguson NM, Laydon D, Nedjati-Gilani G, et al. Impact of non-pharmaceutical interventions (NPIs) to reduce COVID-19 mortality and healthcare demand. Imp Coll COVID-19 Response Team. March 2020:20. doi:https://doi.org/10.25561/77482

6. Tsai TC, Jacobson B, Jha AK. American hospital capacity and projected need for COVID-19 patient care. Health Aff (Millwood). March 2020. doi:10.1377/hblog20200317.457910

7. Petropoulos F, Makridakis S (2020) Forecasting the novel coronavirus COVID-19. PLoS ONE 15(3): e0231236. https://doi.org/10.1371/journal.pone.0231236

8. Du Z, Xu X, Wu Y, Wang L, Cowling BJ, Ancel Meyers L. Serial interval of COVID-19 among publicly reported confirmed cases. Emerg Infect Dis. 2020 Jun [date cited]. https://doi.org/10.3201/eid2606.200357

9. Nunes-Vaz, R., 2020. Visualising the doubling time of COVID-19 allows comparison of the success of containment measures. Global Biosecurity, 1(3), p.None. DOI: http://doi.org/10.31646/gbio.61

10. https://covid19.healthdata.org/projections

11. SAS Institute, version 9.4, Cary, NC

12. https://covid19.colorado.gov/case-data)

